# Automated Online Speech Analytics Reveal Language and Affective Changes in Parkinson’s Disease

**DOI:** 10.1101/2025.11.09.25339865

**Authors:** Fangyuan Cao, Kerrie McAloney, Kishore R. Kumar, Nicholas G. Martin, Puya Gharahkhani, Adam P. Vogel, Miguel E. Renteria

## Abstract

Speech-related changes hold promise as biomarkers in Parkinson’s disease (PD), yet evidence remains preliminary and higher-order language alterations are under-characterised. Leveraging data from 1,168 participants with PD and 541 controls within the Australian Parkinson’s Genetics Study, we report population-level evidence of PD-associated linguistic and affective alterations. Participants with PD exhibited reduced fluency and expressivity, with slower rate, lower reading accuracy, reduced output, simpler syntax, and more neutral, less positive emotional tone. Sex-specific patterns emerged: females used less specific language with fewer nouns and modifiers, while males showed broader vocabulary, higher verb density, and fewer adverbs. Language changes were associated with disease duration and cognitive, mood, and sleep comorbidities, and were linked to reduced communication effectiveness and social engagement. These findings deepen understanding of communicative and affective changes in PD, and our scalable online assessment with automated analytics provide a replicable framework for embedding speech analysis into research and clinical practice.

## Introduction

Parkinson’s disease (PD) is a progressive, multifactorial neurodegenerative disorder that presents with a complex mix of motor and non-motor symptoms. While tremor, rigidity, and bradykinesia are hallmark features, non-motor symptoms such as impaired olfaction, dysarthria, autonomic dysfunctions, cognitive decline, and neuropsychiatric disorders (e.g., sleep and mood disturbances), are increasingly recognised as integral to the disease progression,^1–3^ and, in some cases, more debilitating than motor symptoms.^4,5^ Among these, changes in voice, speech, and cognitive-linguistic function are particularly prevalent and impactful.^5–7^

Up to 90% of people with PD experience speech impairments, typically marked by hypophonia, monotonous prosody, imprecise articulation, and breathy or hoarse voice, collectively resulting in dysarthric speech that compromises intelligibility.^8^ Beyond the voice-speech domain, PD also impacts higher-order linguistic and pragmatic abilities, such as impaired language processing, word-finding difficulties, reduced syntactic complexity, less informative discourse, and blunted affective expressivity.^6,7,9,10^ This constellation of deficits poses a profound communication challenge, contributing to emotional distress, social withdrawal, isolation, and diminished quality of life in people living with PD.^7,11,12^

Speech and language-related alterations often manifest early, sometimes decades before a formal diagnosis, and evolve and deteriorate over time,^13–15^ potentially mirroring underlying progressive neurodegeneration and offering a critical window into the still incompletely defined disease pathophysiology.^1^ Given their high prevalence, early onset, temporal proximity to disease process, speech-related changes also represent a valuable avenue for early diagnosis, intervention planning, and disease progression tracking. A comprehensive understanding of the altered speech and language production in PD is therefore particularly useful not only to tailor more effective, personalised care that addresses the full scope of patient needs but also to advance biomarker discovery.

Speech features have garnered interest as non-invasive, cost-effective biomarkers for early detection, monitoring, and treatment evaluation in PD.^15^ However, despite their promising performance in distinguishing PD from controls, current evidence remains preliminary, with small, constrained cohorts limiting generalisability and impeding clinical translation.^15–17^ Importantly, most existing studies focus narrowly on motor speech features, typically derived from brief, structured speech prompts.^16–18^ In contrast, higher-order language functions are largely absent from clinical and research paradigms, despite their potential to provide unique and complementary insights into the motor, cognitive, and neuropsychiatric facets of PD.^6,9,15,19^ The nature and profile of language production alterations remains underexplored and poorly characterised, with mixed findings across speech rate, syntactic complexity, grammatical accuracy, and verb use.^7,10,13,15,20^

In light of existing research gaps, the present study sought to generate population-level data on linguistic and sentiment characteristics in PD by leveraging a large-scale national cohort from the Australian Parkinson’s Genetics Study (APGS). Specifically, we analysed group-level differences across various linguistic and sentiment features associated with PD, and further examined sex-specific patterns and their relationships with disease characteristics and quality of life metrics using a scalable, data-driven framework.

## Methods

### Study design

This study aimed to investigate linguistic and emotional alterations in the language production associated with PD at the population-level, leveraging cross-sectional data from the APGS. The APGS, led by QIMR Berghofer in Brisbane, Australia, is a large-scale, ongoing initiative established to advance understanding of the aetiological mechanisms and determinants of PD onset and progression by recruiting and following a nationwide cohort of 10,000 individuals with PD and 10,000 controls.^21^ Recognising the prevalence and clinical significance of speech-related symptoms, APGS launched Speech and Parkinson’s, a substudy dedicated to comprehensively characterising speech and language impairments in PD and identifying speech-based biomarkers to inform diagnosis, monitoring, and clinical management, providing the cohort for the present analysis.

Ethics approval was granted by the QIMR Berghofer Human Research Ethics Committee (P3711), with a protocol amendment approving the Speech and Parkinson’s substudy (P2210). All participants provided informed consent. Data are securely stored and managed in accordance with the Commonwealth Privacy Act (1988) and National Health and Medical Research Council (NHMRC) Guidelines.

### Study population

Recruitment procedures of the APGS have been described previously.^21^ Briefly, the APGS employs a combination of assisted mailouts, national campaigns, public appeals, and referrals. Full-scale nationwide recruitment was launched in April 2022. Cases included individuals residing in Australia with a confirmed PD diagnosis or ongoing treatment (e.g., Levodopa). Controls comprised individuals aged ≥45 years, residing in Australia, with no personal or family history of PD. Participants completed a structured questionnaire, online or paper-based, and provided a saliva sample for genotyping.

From August 2024 onwards, APGS participants who had completed the questionnaire and consented to future contact were invited to the Speech and Parkinson’s substudy. The present study included 1,168 participants with PD and 541 unaffected controls recruited by 31 May 2025. An overview of the study design and participant enrolment is shown in Figure 1.

**Figure 1.**
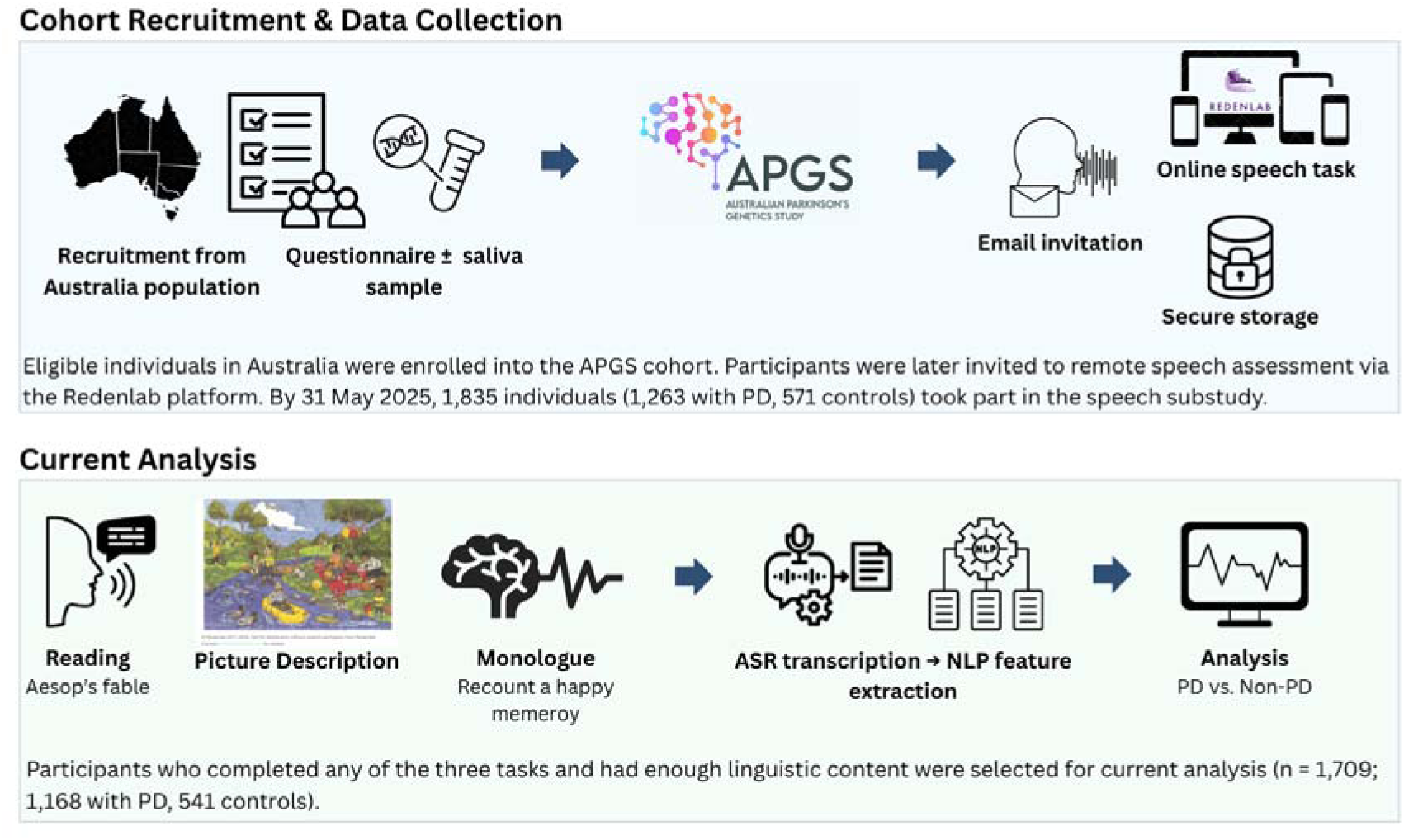
**Overview of study design and data processing pipeline**

### Speech task and Dysarthria Impact Scale

Participants completed a set of speech tasks online via the Redenlab® Ltd. platform (https://redenlab.com/), a validated system accessible on various digital devices, including smartphones, tablets or computers. The speech battery comprised seven tasks designed to capture both structured and spontaneous speech, namely: sustained phonation of the vowel /a/, syllable repetition of /pa//ta//ka/ as quickly and clearly as possible for 5 seconds, reading a phonetically balanced paragraph (Aesop’s fable, “The north wind and the sun”), description of a cartoon picture (“Fishing”; https://redenlab.com/picture-description-stimuli/), a monologue recounting a happy memory for 1-2 minutes, and semantic fluency (animal naming) and phonemic fluency tasks (‘f’ words naming).^22^ In the current study, we analysed the reading, picture description, and monologue tasks.

Participants also completed the Dysarthria Impact Scale (DIS-6),^12^ a six-item standardised tool assessing patient-perceived impact of their speech-related impairment on functional communication, psychosocial well-being, and overall quality of life. The DIS-6 comprises six items that address aspects of communication and its consequences: Q1) My speech affects my social life; Q2) I feel isolated or lonely because of my speech; Q3) People unfamiliar with me have difficulty understanding my speech; Q4) Because of my speech I avoid talking on the phone; Q5) Because of my speech it is difficult for me to describe events; and, Q6) Because of my speech I have difficulty participating in fast or complex conversations. Each item was rated on a five-point Likert scale: fully agree, agree, neither agree nor disagree, do not agree, and do not agree at all. Lower scores represent higher burden of impairment.

### Feature extraction

Audio recordings were transcribed to texts using OpenAI’s Whisper model for English.^23^ The medium model was selected for its optimal balance of transcription accuracy, performance stability, and computational efficiency, following a comparative evaluation of the base, small, medium, and large model variants on a subset of the sample. Punctuation in the picture description and monologue transcripts was removed and restored using the Python library *DeepMultilingualPunctuation*, which improves accuracy for spoken text.^24^

For all three tasks, we computed transcript duration (the time interval between the onset of the first word and the offset of the final word), total words produced, and speaking rate (total words / transcript duration). Read-aloud accuracy was proxied using word error rate (WER), which captured both participant-generated errors and potential automatic speech recognition (ASR) transcription errors, which may arise from reduced articulation clarity (e.g., in PD) or ASR autocorrection. Punctuation was removed prior to WER computation to focus on the core lexical content.

Lexical and syntactic complexity were evaluated using a range of sentence- and phrase-level features extracted from the picture description and monologue tasks with *Stanza*, a Python-based natural language processing (NLP) toolkit developed by Stanford for advanced linguistic analysis.^25^ *Stanza* was selected over other NLP libraries for its research-grade accuracy, nuanced handling of linguistic structures, and robust syntactic parsing capabilities.^25^ Sentence-level measures included the number of T-units (an independent clause plus its subordinate clauses), mean length of T-units (MLT), and the proportion of subordinate clauses. Phrase-level features captured frequencies of lexical categories (nouns, pronouns, verbs, auxiliaries, adjectives, adverbs, interjections, noun- and verb-group modifiers), ratios of nouns to pronouns and verbs to auxiliaries, content density, and lexical diversity (type-token ratio, TTR; see Table 1 for feature definitions and rationale).

**Table 1.**
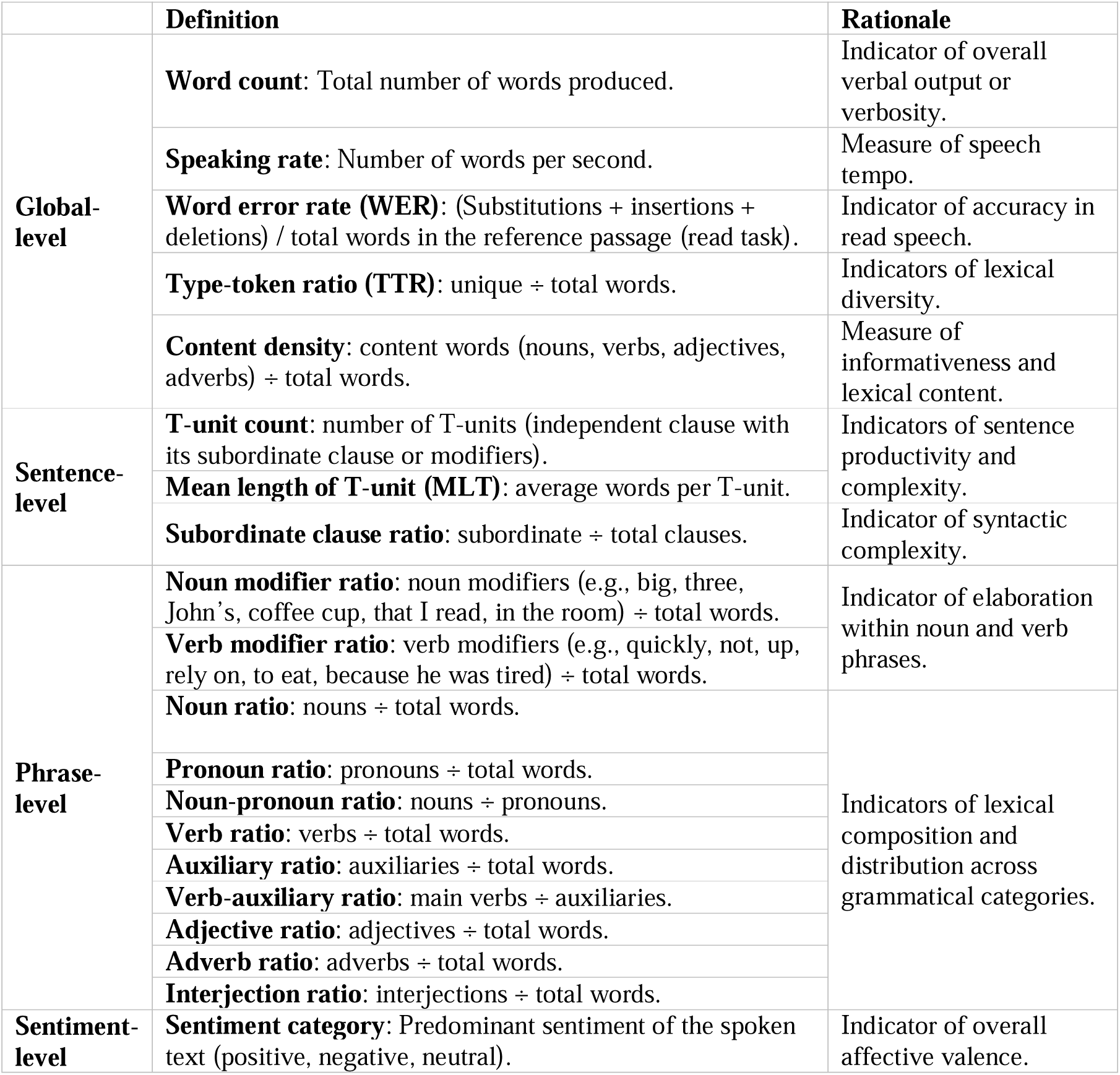
Definition and rationale of measured linguistic variables.

We explored the potential influence of PD on emotional valence by performing sentiment analysis on transcripts from the picture description and monologue tasks using the RoBERTa model from the Hugging Face *Transformers* library.^26^ This model was selected for its high accuracy and ability to capture nuanced emotional content in naturalistic, informal language.^26^ The analysis generated a probability score (ranging from 0 to 1) for each sentiment category: positive, negative, or neutral, along with a categorical label corresponding to the highest-probability sentiment.

Duplicated recordings were addressed using a two-step procedure. First, recordings were excluded if they contained fewer than 90 words for the reading task or fewer than 50 words for the picture description and monologue tasks. These criteria account for instances where participants may have been interrupted during an initial attempt and later started a new recording. Second, for any remaining duplicates, the assessed measures were averaged to yield a single representative value per variable for each participant.

To uphold analytical validity, only participants with sufficient verbal content were retained for analysis: transcripts of reading task required ≥90 words, and picture description and monologue transcripts required ≥40 words. For the reading task, the 90-word cut-off, corresponding to 80% task completion, was selected to balance transcript informativeness and sample retention, following a comparison of different thresholds (80%-90% task completion). The final sample included participants with a valid transcript from at least one task.

### Statistical analysis

The primary analyses assessed the impact of PD on linguistic and sentiment outcomes. Linear regression was used for continuous linguistic measures, and multinomial logistic regression for sentiment classifications. Potential confounders were evaluated and adjusted using a stepwise approach: an unadjusted model assessed baseline group differences (PD vs. non-PD); Model 1 adjusted for age and sex; Model 2 additionally included education and multilingualism.

Normality of continuous outcomes was assessed by visually inspecting histograms and quantile-quantile plots, with appropriate transformations (e.g., log-transformation) applied when assumptions were violated. Multiple testing was controlled by applying the Benjamini-Hochberg false discovery rate (FDR) correction across all assessed outcomes, with FDR-adjusted *p* < 0.05 deemed significant.

Acknowledging sex-related variations in PD presentation^27,28^ and language use,^29^ we conducted within- and between-sex comparisons using the same regression-based approach. Specifically, we examined four pairings: 1) male cases vs. male controls, 2) female cases vs. female controls, 3) male cases vs. female cases, and 4) female controls vs. male controls. To estimate all four pairwise contrasts, models were re-run iteratively with each group serving as the reference category. FDR correction was applied across all comparisons and outcomes. All regression models were adjusted for age, education, and multilingualism.

To gain deeper insights into language and affective expression, we further explored clinical correlates of linguistic and sentiment outcomes within the PD group. Clinical variables included age of onset, early-versus late-onset PD (age of onset <50 or ≥50 years), age at diagnosis, Levodopa use, sleep complaints (e.g., difficulty falling asleep, vivid or frightening dreams), and self-reported comorbidities across four domains: sleep, cognitive, and mood disorders, as well as chronic pain. Sleep disturbances encompassed self-reported symptoms, such as difficulty falling or staying asleep, excessive daytime sleepiness, vivid or frightening dreams, sleep talking or movement, unpleasant leg sensations, and snoring. Sleep comorbidities included diagnosis of rapid eye movement sleep behaviour disorder (RBD), restless legs syndrome, sleep apnoea, narcolepsy, and insomnia. Cognitive disorders comprised mild cognitive impairment, Alzheimer’s disease, Parkinson’s disease dementia, frontotemporal dementia, vascular dementia, dementia with Lewy bodies, and other forms of dementia. Mood disorders included depression, anxiety, and bipolar disorder. Chronic pain was defined as pain persisting for more than three months.

We also examined relationships between linguistic patterns and self-perceived quality of life and social impact using logistic regression, adjusting for the covariates aforementioned. Responses on the DIS-6 were dichotomised into ‘agree’ (combining ‘fully agree’ and ‘agree’) and ‘disagree’ (all other categories). To minimise multicollinearity, Principal Component Analyses were applied to two linguistic domains: speech quantity (total words, T-units, MLT, TTR, and content density) and lexical composition (ratios of nouns, pronouns, verbs, auxiliaries, their syntactic modifiers), with the first principal component (PC1) for each domain used as a composite score in regression models. For the reading task, models included speaking rate, WER, and covariates. For the picture description and monologue tasks, models included speaking rate, the two PC1 scores, syntactic complexity (subordinate clause ratio), task type, along with the covariates.

Finally, we conducted an exploratory correlation analysis among linguistic features using Spearman rank correlation to help interpret group-level differences and gain insight into structure of language production.

### Sensitivity analysis

Baseline characteristics were compared between participants who passed versus failed quality control for analysis, and participants who responded to the substudy (responders) with those who did not (non-responders) to evaluate potential selection bias.

To gauge the potential impact of ASR model performance on downstream analyses, FC manually curated audio recordings from a 10% random subset of participants, including 108 cases and 46 controls, enrolled by 30 April 2025, as participants recruited in May 2025 were added to the analysis later. This subset was then analysed separately and compared with results derived from automatic transcripts.

All statistical analyses and visualisations were conducted in Python (3.12.5) using the *Pandas*, *NumPy*, *Scipy*, and *Matplotlib* libraries and R (4.4.1) with the *tidyverse* and *ggplot2* packages.

## Results

### Study sample

As of 31 May 2025, 1,835 participants completed the online speech assessment. Participants with missing demographic data (n = 1) or without recordings for any of the three assessed tasks (n = 80) were excluded. An additional 45 participants were excluded due to insufficient linguistic content. The final analysis included 1,709 participants (1,168 with PD and 541 without), of whom >93% were of European ancestry (Table 2).

**Table 2.**
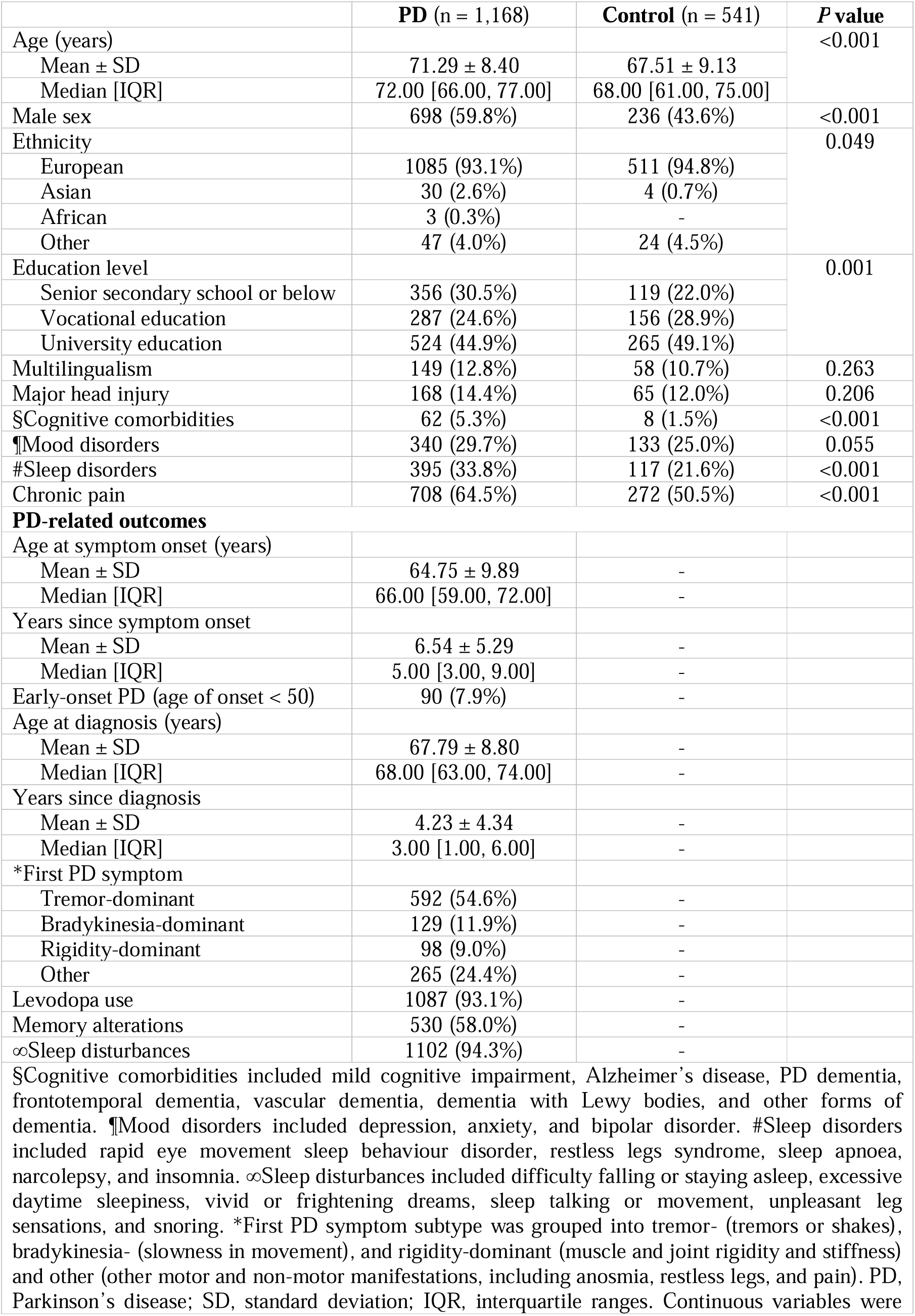
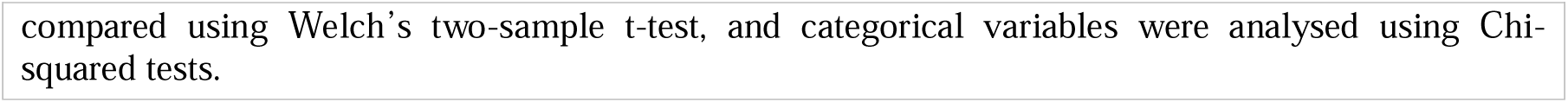
Baseline characteristics of the study sample.

Compared with controls, the PD group was older (71.3 ± 8.4 vs. 67.5 ± 9.1 years, *p* < 0.001), more often male (60% vs. 44%, *p* < 0.001), less likely to have tertiary education (70% vs. 78%, *p* = 0.001), and showed higher prevalence of cognitive (5% vs. 2%, *p* < 0.001) and sleep comorbidities (34% vs. 22%, *p* < 0.001), as well as chronic pain (65% vs. 51%, *p* < 0.001). Among participants with PD, the mean disease duration was 6.5 ± 5.3 years from symptom onset and 4.2 ± 4.3 years from diagnosis.

### Group-level differences

Of the nineteen linguistic features examined, sixteen, along with language sentiment, differed significantly between PD and control groups (Figures 2-3; see Supplementary Data, Table S1-S2 for full details).

**Figure 2.**
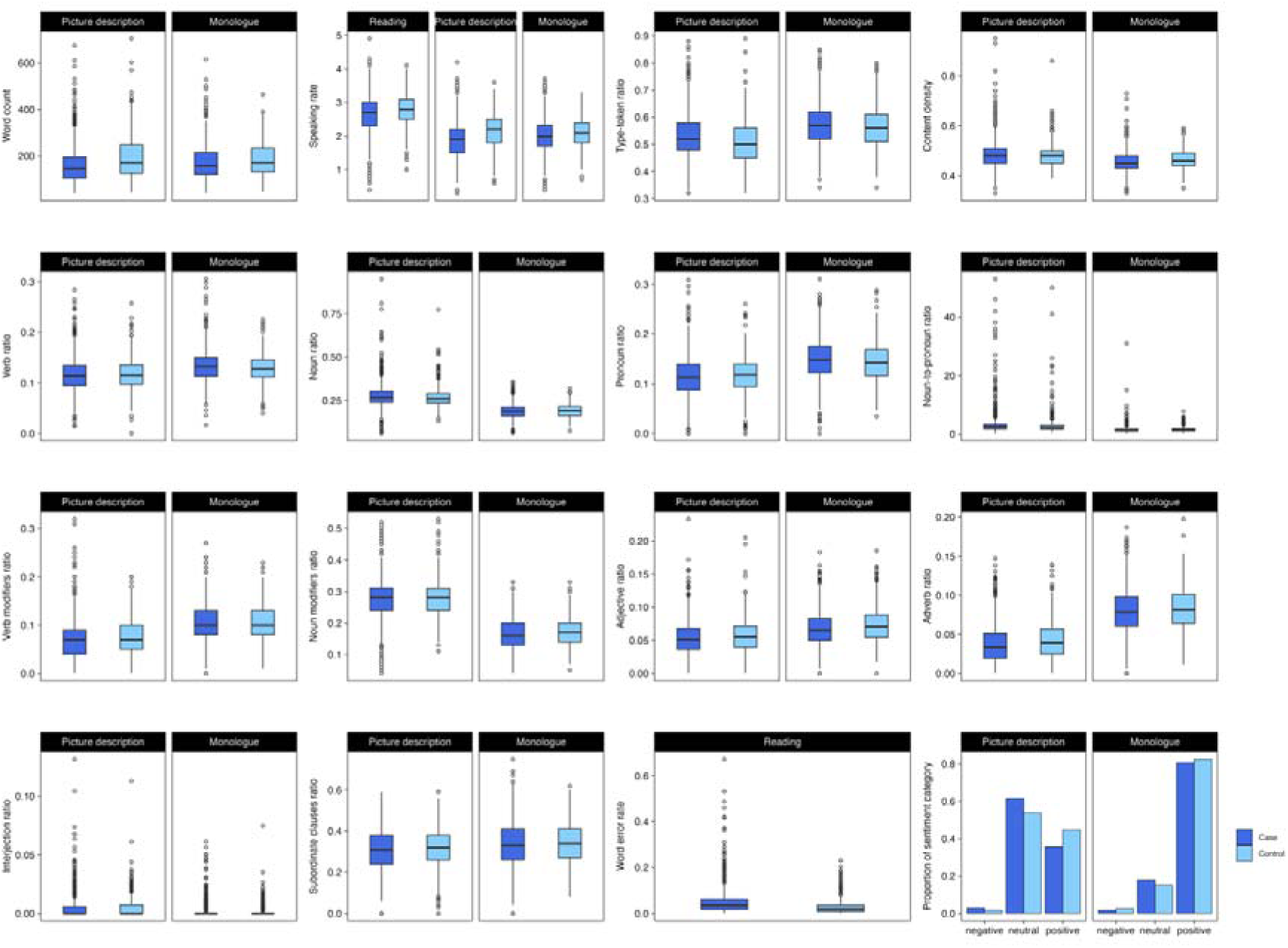
Linguistic and sentiment characteristics in participants with Parkinson’s disease and unaffected controls. The boxplots illustrate the distribution of features with significant group differences between participants with Parkinson’s disease and unaffected controls across the reading, picture description, and monologue tasks.

**Figure 3.**
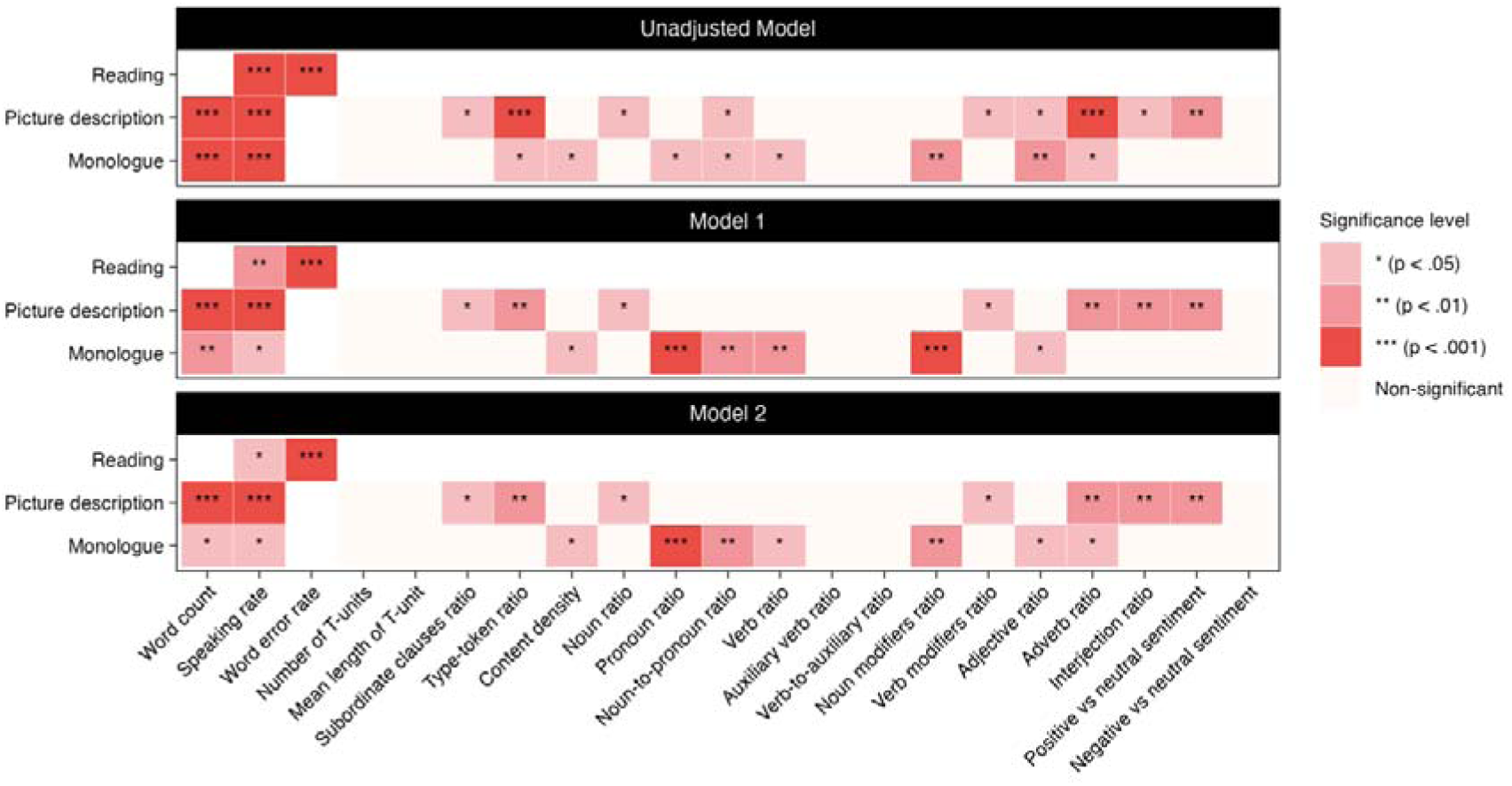
Differences in linguistic and sentiment characteristics between groups, across covariate adjustment. The heatmap displays results from case-controls comparisons for the reading, picture description, and monologue tasks across three regression models: the unadjusted model, Model 1 (adjusted for age and sex), and Model 2 (further adjusted for multilingualism and educational level). False discovery rate (FDR) correction was applied across all assessed outcomes.

In the reading task, participants with PD exhibited slower speaking rates (*p* < 0.001) and more frequent reading errors (higher WER, *p* < 0.001) compared to controls, patterns that persisted across covariate adjustments.

The picture description task revealed group differences in syntactic, lexical, and expressive patterns. Participants with PD produced fewer words (*p* < 0.001), spoke more slowly (*p* < 0.001), used fewer subclauses (*p* = 0.037), verb modifiers (*p* = 0.037), adverbs (*p* < 0.001), and interjections (*p* = 0.012), and conveyed less positive language (*p* = 0.002) than controls. In contrast, noun ratio (*p* = 0.044) and vocabulary diversity (TTR, *p* < 0.001) were slightly increased in the PD group. These differences remained significant after covariate adjustment, whereas the observed reduction in adjective use (*p* = 0.037) was no longer significant after adjusting for age and sex.

Consistent with the picture description, participants with PD spoke more slowly (*p* < 0.001) and produced fewer words (*p* < 0.001) in the monologue task. Additional lexical differences included greater reliance on pronouns over specific nouns (higher pronoun ratio, *p* = 0.011; lower noun-to-pronoun ratio, *p* = 0.037), reduced use of adjectives (*p* = 0.001) and noun modifiers (*p* = 0.008), higher verb ratio (*p* = 0.014), and a shift toward function words over content words, as reflected by lower content density (*p* = 0.015). These patterns persisted across all covariate-adjusted models. Reductions in adverb use (*p* = 0.012) and more diverse vocabulary (*p* = 0.011) were noted but were no longer significant after adjusting for age and sex.

### Sex-based differences

Within sex groups, both male and female participants with PD exhibited higher reading error rates than their respective controls in the reading task (*p* < 0.001), while slower speaking rates were observed specifically in males (*p* = 0.044) (Figure 4; see Supplementary Data, Table S3 and Figure S1 for full details). In the picture description task, word production (*p* < 0.001 for males; *p* = 0.019 for females) and speaking rate (*p* < 0.001 for males; *p* = 0.004 for females) were reduced in both sexes with PD, with more pronounced reductions in males. Male participants with PD used less expressive language, particularly fewer adverbs (*p* = 0.006) and interjections (*p* = 0.001), but showed slightly broader vocabulary (*p* = 0.008) and a higher noun-to-pronoun ratio (*p* = 0.025) than male controls. In the monologue task, males with PD again spoke more slowly (*p* = 0.031) and, despite higher verb density (*p* = 0.002), used less adverbs (*p* = 0.030). Female participants with PD relied more on pronouns (lower noun-to-pronoun ratio, *p* = 0.006) and produced fewer noun modifiers (*p* = 0.001) and lower content density (*p* = 0.007) compared to female controls.

**Figure 4.**
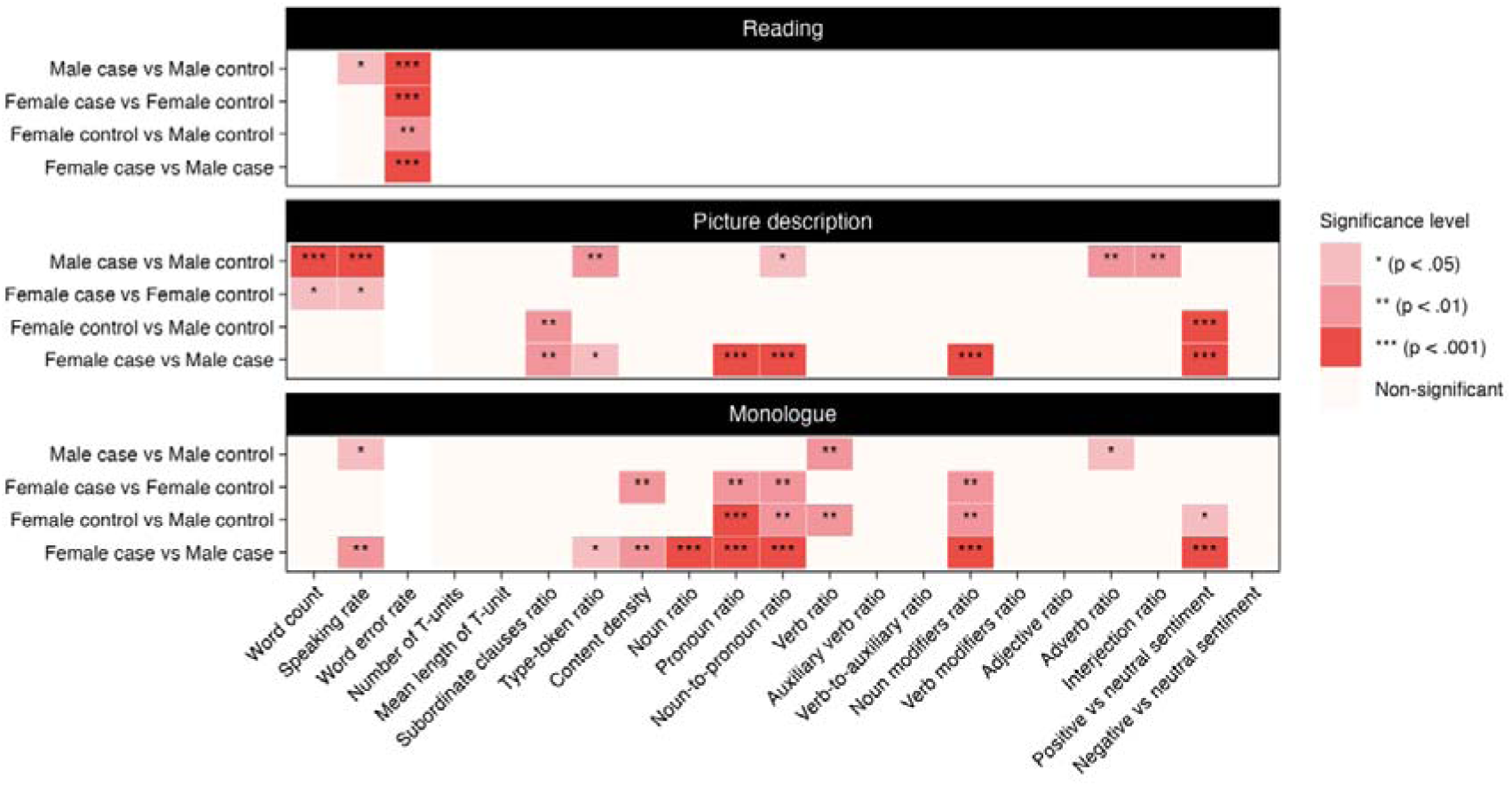
Within and between sex group differences in linguistic and sentiment characteristics. The heatmap displays results from group comparisons within and between males and females across three tasks. All models adjusted for age, education, and multilingualism. False discovery rate (FDR) correction was applied across all comparisons and outcomes.

When comparing sexes, regardless of PD status, males consistently performed worse on the reading task (*p* = 0.002 for controls; *p* < 0.001 for PD group) and showed a trend toward less positive, more neutral sentiment in the picture description (*p* < 0.001 for both PD and control groups) and monologue tasks (*p* = 0.023 for controls; *p* < 0.001 for PD group), whereas females used fewer subordinate clauses in picture description (*p* = 0.007 for controls; *p* = 0.003 for PD group). Within the PD group, males spoke more slowly during the monologue (*p* = 0.006) but exhibited greater lexical diversity in picture description (*p* = 0.032) and monologue (*p* = 0.047), while females showed higher use of pronouns and lower noun modifier use in both tasks (*p* < 0.001 for both). During the monologue, females with PD also demonstrated reduced content density (*p* = 0.006) and lower noun ratios (*p* < 0.001), with similar patterns of higher pronoun ratios (*p* < 0.001) and lower noun-to-pronoun ratios (*p* = 0.002) observed among controls.

### Correlation with clinical factors

Longer disease duration, measured from symptom onset or diagnosis, was consistently associated with increased reading errors and slower speaking rates during both reading and picture description (Table 3). Extended disease duration was also linked to more frequent use of interjections and less use of subclauses, as well as reduced positive emotional expression.

**Table 3.**
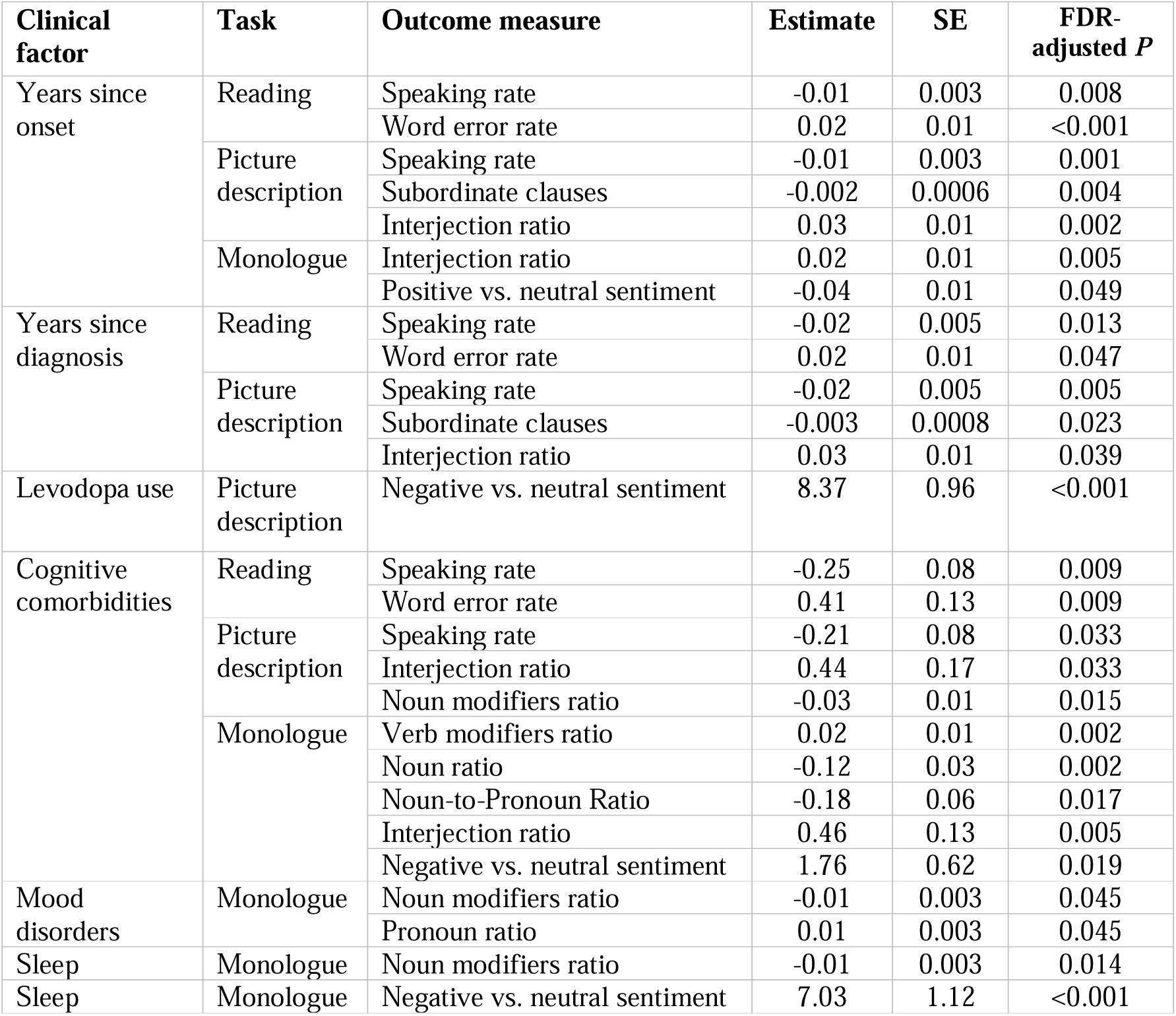
Significant associations between clinical factors and linguistic outcomes within the Parkinson’s disease group.

Cognitive comorbidities were associated with slower, more error-prone reading, reduced use of specific nouns, increased interjections and verb modifiers, and a shift toward more negative sentiment. Mood-related comorbidities were linked to increased pronoun use and fewer noun modifiers during the monologue, while sleep disorders were associated with reduced noun modifier use. Sleep disturbances and Levodopa use were related to greater negative sentiment. No associations were observed for early-versus late-onset PD or chronic pain.

### Correlation with self-perceived impact

Participants with PD reported a greater burden across all DIS domains compared to controls (Figure 5). Specifically, 59% of participants with PD felt their speech affected their social life versus 40% of controls, and 17% reported feelings of isolation versus 4% of controls. Difficulties with being understood (28% vs. 4%), phone conversations (18% vs. 4%), event description (33% vs. 4%), and engaging in complex conversations (46% vs. 7%) were also substantially higher in the PD group, with males experiencing more pronounced effects than females (see Supplementary Data, Table S4 and Figure S2).

**Figure 5.**
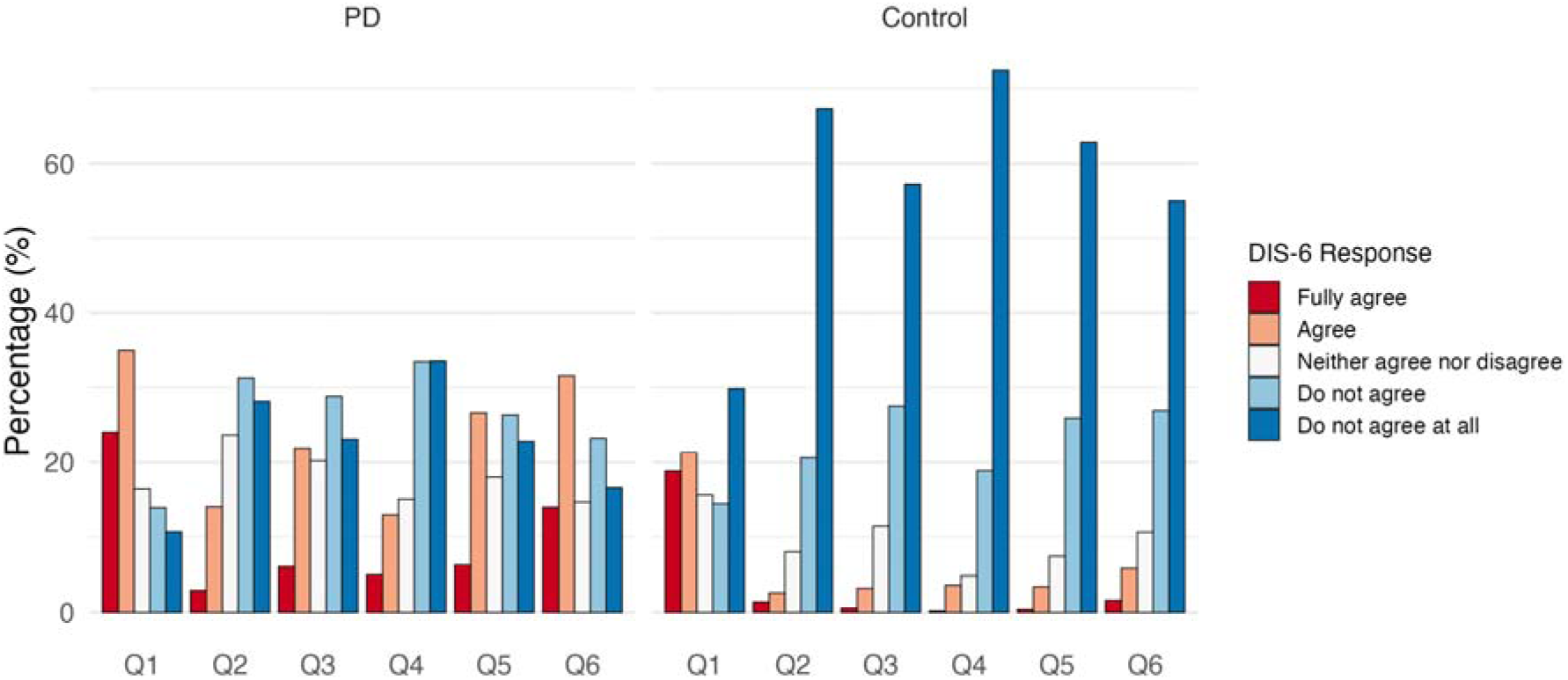
Participant response to Dysarthria Impact Scale (DIS-6) The bar chart illustrates the responses to the Dysarthria Impact Scale (DIS-6) in participants with Parkinson’s disease and controls. The questions asked were: Q1) My speech affects my social life; Q2) I feel isolated or lonely because of my speech; Q3) People unfamiliar with me have difficulty understanding my speech; Q4) Because of my speech I avoid talking on the phone; Q5) Because of my speech it is difficult for me to describe events; and Q6) Because of my speech I have difficulty participating in fast or complex conversations.

For read speech, higher WER was associated with greater self-perceived impact across all DIS items. Slower speaking rate correlated with increased interference in social engagement, reduced intelligibility, and challenges in describing events or participating in fast or complex conversations. Across narrative speech tasks (picture description and monologue), slower speaking rate correlated with greater perceived impact on all DIS items. Lexical features were associated with difficulties in phone communication, social withdrawal, event description, and complex conversations, while syntactic simplification was linked to greater difficulty describing events (see Supplementary Data, Table S5 for full details).

### Feature interrelationship

Interrelationships were observed among linguistic features. Longer responses in general were characterised by faster speech and a higher number of T-units: word count was positively correlated with speaking rate and T-units count (r = 0.61 and 0.66, respectively), with a trade-off between the quantity and length of T-units (r = –0.51). Vocabulary richness was inversely related to speaking rate (r = –0.46), word count (r = –0.71), and T-units count (r = –0.45). Noun-related features (noun ratio, noun-to-pronoun ratio, and noun modifiers ratio) were strongly intercorrelated (r > 0.83), whereas verb-related features showed only moderate intercorrelations (r = 0.30-0.58) and were negatively associated with noun-based features (r = –0.38 to –0.63; see Supplementary Data, Figure S3).

### Sensitivity analysis

Due to insufficient audio content, we excluded 194 (11.9%), 57 (3.7%), and 25 (1.6%) participants from the study sample for the reading, picture description, and monologue tasks, respectively. The distribution of most demographic and PD-related variables was comparable between participants retained and those excluded. However, some differences were observed: participants retained in the analysis were, on average, younger (68-69 vs. 70-73 years), have higher education (46-48% with university degrees vs. 24-34%), and were less likely to have cognitive comorbidities (4% vs. 9% for the reading task; see Supplementary Data, Table S6).

Participants who responded to the substudy, compared to non-responders, were slightly younger (69 vs. 70 years), more likely to be female (45% vs. 41%), and more highly educated (46% with university degrees vs. 36%), and had fewer cognitive comorbidities (4% vs. 8%) and mood disorders (28% vs. 34%). They were also less likely to have a PD diagnosis (69% vs. 75%) and, among those with PD, had a shorter disease duration (5.2 vs. 6.8 years since onset; 3.3 vs. 4.8 years since diagnosis; see Supplementary Data, Table S7).

Group comparisons using manually curated transcripts from the random subset demonstrated consistent differences in linguistic and sentiment features between participants with PD and those without, supporting the reliability of the ASR model in this study (see Supplementary Data, Table S8 for demographic characteristics of the subset and Table S9 for analysis results).

## Discussion

This study, integrating a population-based cohort with an automated analytic pipeline, provides evidence of distinct linguistic and emotional language patterns in PD across speech contexts. Affected individuals consistently displayed slower, less fluent, shorter, less complex, and emotionally blunted speech. Sex-based differences emerged in noun/verb phrase use and expressive features. Longer disease duration and comorbidities were associated with increased reading errors, slower speech, and diminished expressiveness. Individuals with PD, particularly males, reported markedly greater everyday communication challenges, linked to specific linguistic features.

A key finding was the reduced reading accuracy in PD compared to controls, which was more pronounced in males and associated with longer disease duration and cognitive comorbidities. The reading errors likely reflect combined motor and cognitive-linguistic disruptions. Progressive bradykinesia and rigidity of the speech musculature and oculomotor dysfunction compromise articulatory precision and fluency,^30^ while cognitive decline in working memory, language processing, and speech planning further limits the ability to decode and verbalise written text efficiently.^31^ The more severe motor and cognitive deterioration often observed in males^27,28^ may explain their poorer performance. Automated analyses suggest that reading errors and disfluencies may predict cognitive decline, although the lack of control groups has limited conclusions about PD-specific effects.^32^ Our study extends previous work by demonstrating measurable deficits in read speech among individuals with PD.

We observed a consistent slowdown in connected speech, which becoming more pronounced with advancing PD and cognitive comorbidities. Findings on speech rate abnormalities in PD have been mixed: some show reduced rates that decline over time, others report increased or unchanged rates, ^20,33–36^ likely due to small, heterogeneous samples and differing task demands. By leveraging a population-based cohort and multiple elicitation tasks, our results indicate a uniform speech slowdown across contexts, potentially reflecting generalised physiological alterations in speech motor control associated with basal ganglia dysfunction,^37^ effects not solely driven by task-specific cognitive or linguistic demands.

Distinct syntactic and lexical changes in PD were observed. At the sentence level, participants with PD produced shorter, less informative utterances and simplified syntax with fewer clauses than controls during narrative tasks. Motor limitations and cognitive constraints, including executive slowing, memory deficits, and language impairments that hinder effective information organisation and delivery may underlie reduced verbosity and information density.^9,38,39^ Evidence on syntactic and grammatical impairments in PD is mixed, with most studies focusing on comprehension rather than production.^7,9^ Our results indicate syntax simplification in PD, with effects varying by task: it was evident in picture description but not in monologue. Compared to free-form monologue, picture description imposes additional linguistic and conceptual demands, such as visual processing, lexical retrieval, and mapping observations onto structured sentences, which may prompt syntax simplification to preserve essential communication. This pattern aligns with broader cognitive constraints on organisation, planning, and abstract thought linked to frontal lobe function, as well as short-term memory and attention,^40^ which are increasingly taxed as structural demands rise.

At the phrase level, participants with PD exhibited reduced lexical specificity and elaboration, with distinct sex-specific profiles: females relied more on pronouns and used fewer noun modifiers, whereas males showed higher verb density and lexical diversity but reduced adverb use. These differences may reflect differential motor or cognitive involvement. Males generally experience greater motor-speech impairment,^15,27^ which may lead to broader or more effortful lexical selection, resulting in more content-dense words in shorter utterances to maintain communicative effectiveness. Supporting this, we noted a negative correlation between speech and sentence length and vocabulary richness. In contrast, females’ tendency to use self-referential language (substitution of specific words with more general forms) may reflect reduced lexical retrieval efficiency and differing communication priorities shaped by gendered language patterns.^29^ These findings underscore the need to examine sex-specific language patterns in PD to refine language-based biomarkers and understand disease-related cognitive mechanisms.

Verb- and noun-related features have been identified as key discriminators between PD and controls in machine learning studies, reinforcing PD’s influence on lexical patterns.^41^ However, findings are inconsistent: while selective verb deficits, particularly in action-verb naming, are often reported,^7,41^ spontaneous speech may show increased verb use and reduced nouns and fillers.^42^ These discrepancies underscore the need for systematic analyses across speech contexts, as sentence generation fundamentally differs from single-word production, requiring integration of multiple words’ meanings and grammatical roles.^9^ Consistent with this, our results revealed that lexical changes in PD were linked to cognitive, mood, and sleep comorbidities, highlighting a complex interplay of linguistic, cognitive, and affective factors in shaping language use.

We also observed emotional flattening in participants with PD, particularly during picture description tasks with no explicit emotional cues. This aligns with prior evidence of impaired emotion recognition, blunted vocal, facial, and bodily expressions, and affective and mood dysfunctions in PD, phenomena linked to dopamine depletion brain regions (e.g., amygdala, orbitofrontal cortex, basal ganglia) critical for emotion and social cognition.^43,44^ Longer disease duration was associated with more neutral and less positive language sentiment, suggesting progressive emotional dysfunction from cumulative neurodegenerative changes. Levodopa use correlated with increased negative sentiment, potentially reflecting neuropsychiatric side effects from mesolimbic dopaminergic overstimulation. These findings underscore the complex interplay of neurobiological and psychological factors in PD and the need to better understand emotional dysfunction to inform targeted, holistic interventions.

Finally, participants with PD, particularly males, reported substantial communication difficulties compared to controls, and specific linguistic patterns were associated with greater challenges in social communication, intelligibility, event description, and participation in complex conversations. These findings indicate that such features may serve as targets for clinical assessment and therapeutic intervention to improve communication outcomes.

To our knowledge, this is the first population-level investigation to comprehensively characterise language and sentiment expression in PD. By analysing diverse linguistic data across multiple natural speech contexts and linking them to sex, clinical features, and quality of life, we reveal nuanced language patterns and clinically relevant associations beyond previously reported group-level differences. Our cohort of >1,700 participants enabled detection of subtle effects and robust, generalisable insights into the linguistic and affective manifestations of PD. Finally, our scalable analytic framework offers an ecologically valid, practical model for future longitudinal and cross-national research.

We acknowledge several limitations. First, our remote recruitment and data collection, while enabling broader participant reach, introduces variability in recording environments and precluded control over medication state at the time of recording. We assumed most participants were in the “on” state, allowing them to complete the tasks. Despite reduced experimental control, our approach demonstrates the feasibility of remote assessment, overcoming geographic, mobility, and scheduling barriers compared to in-clinic evaluations. The remote design also limits detailed clinical phenotyping (e.g., symptom severity); consequently, we used years since onset and diagnosis as approximate indicators of disease stage. Second, the observed sociodemographic and PD-related differences between respondents and non-respondents raise potential for selection bias. To help mitigate this, reminder messages to non-respondents are planned. Lastly, while our automated analytical pipeline is scalable and resource-efficient, it may be less accurate than human annotation, especially for dysarthric speech where model performance can degrade. Nevertheless, manual review of a subset suggested minimal impact on our results. Our future work will integrate acoustic, linguistic, demographic, clinical, and genetic data to better characterise PD symptomatology and improve monitoring and management.

In conclusion, this study represents the first large-scale, population-based investigation of linguistic and affective changes in PD. Our findings reveal novel patterns of language alterations, sex-based differences, and clinical correlates, shedding light on their nature and clinical significance. The scalable and adaptable methodological framework we presented establishes a replicable foundation for future efforts to incorporate speech analysis into clinical assessment and monitoring of neurodegenerative disease.

## Data availability

The original audio recordings supporting this study are not publicly available, as they contain information that could compromise participant privacy. However, anonymised data underlying the study findings may be made available to qualified researchers upon reasonable request to the corresponding authors. Data sharing will be contingent upon obtaining all necessary ethical approvals and completion of an institutional data transfer agreement. Researchers may contact the corresponding authors to request access to the code used for data processing and analysis.

## Code availability

Researchers may contact the corresponding authors to request access to the code used for data processing and analysis.

## Author contributions

FC conceptualised the study, conducted speech processing and transcription, performed statistical analyses, and drafted the manuscript. KM oversaw study operations, including participant engagement and the implementation of the Speech and Parkinson’s substudy. APV, MER, NGM, and KRK contributed to data interpretation, provided analytical input, and critically reviewed the manuscript. PG advised and reviewed the manuscript. APV and MER supervised the study and led the manuscript revision. All authors reviewed and approved the final version of the manuscript for submission.

## Acknowledgements

The authors gratefully acknowledge the invaluable contributions and commitment of the participants in the APGS, without whom this research would not have been possible. This project was supported by the Global Parkinson’s Genetics Program (GP2). GP2 is funded by the Aligning Science Across Parkinson’s (ASAP) initiative and implemented by The Michael J. Fox Foundation for Parkinson’s Research (https://gp2.org). For a complete list of GP2 members see https://gp2.org.

## Supplementary information

Supplementary Data Figures

Supplementary Data Tables

