## Supplementary Figures for "Automated Online Speech Analytics Reveal Language and Affective Changes in Parkinson’s Disease"

### Supplementary Information

#### Supplementary Figures

##### Figure S1 Linguistic and sentiment characteristics in individuals with PD and unaffected controls, stratified by sex

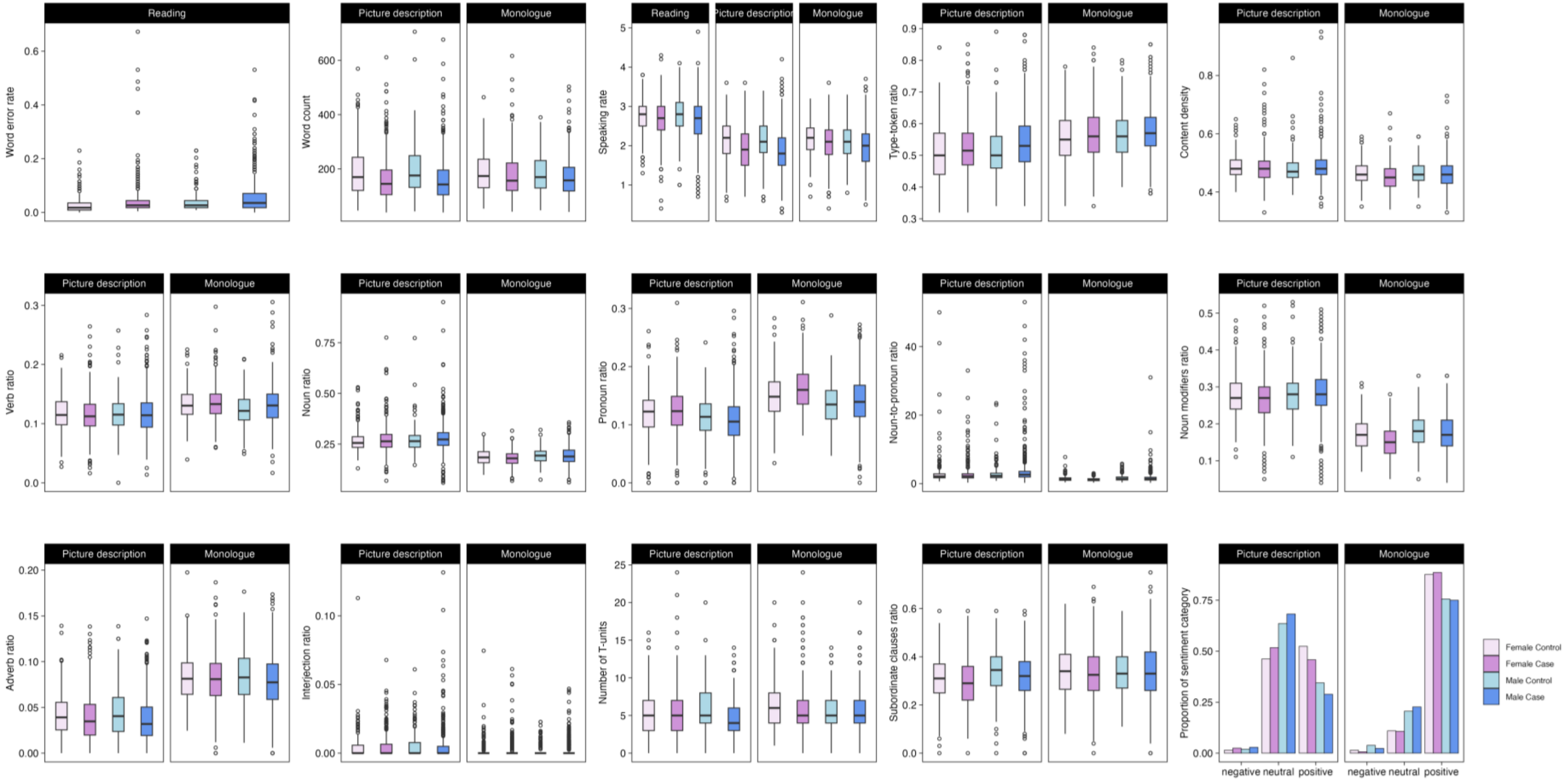


The boxplots illustrate the distribution of features with significant group differences between participants with PD and unaffected controls, stratified by sex, across the reading, picture description, and monologue tasks.

**Figure S2 Response to Dysarthria Impact Scale, stratified by sex**

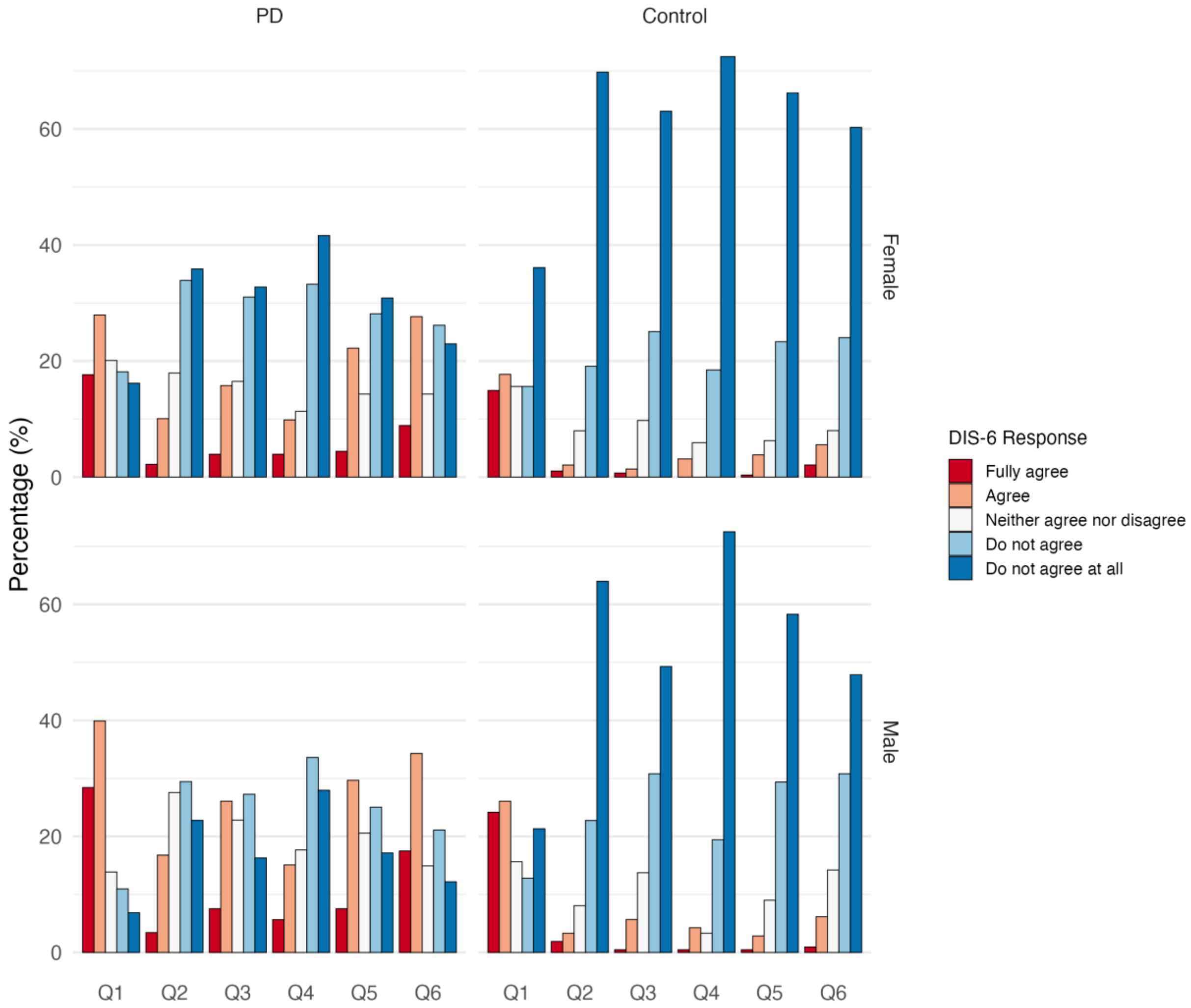


The bar chart illustrates the responses to the Dysarthria Impact Scale in female and male participants with and without PD. The questions asked were: Q1) My speech affects my social life; Q2) I feel isolated or lonely because of my speech; Q3) People unfamiliar with me have difficulty understanding my speech; Q4) Because of my speech I avoid talking on the phone; Q5) Because of my speech it is difficult for me to describe events; and Q6) Because of my speech I have difficulty participating in fast or complex conversations.

**Figure S3 Correlation among linguistic features**

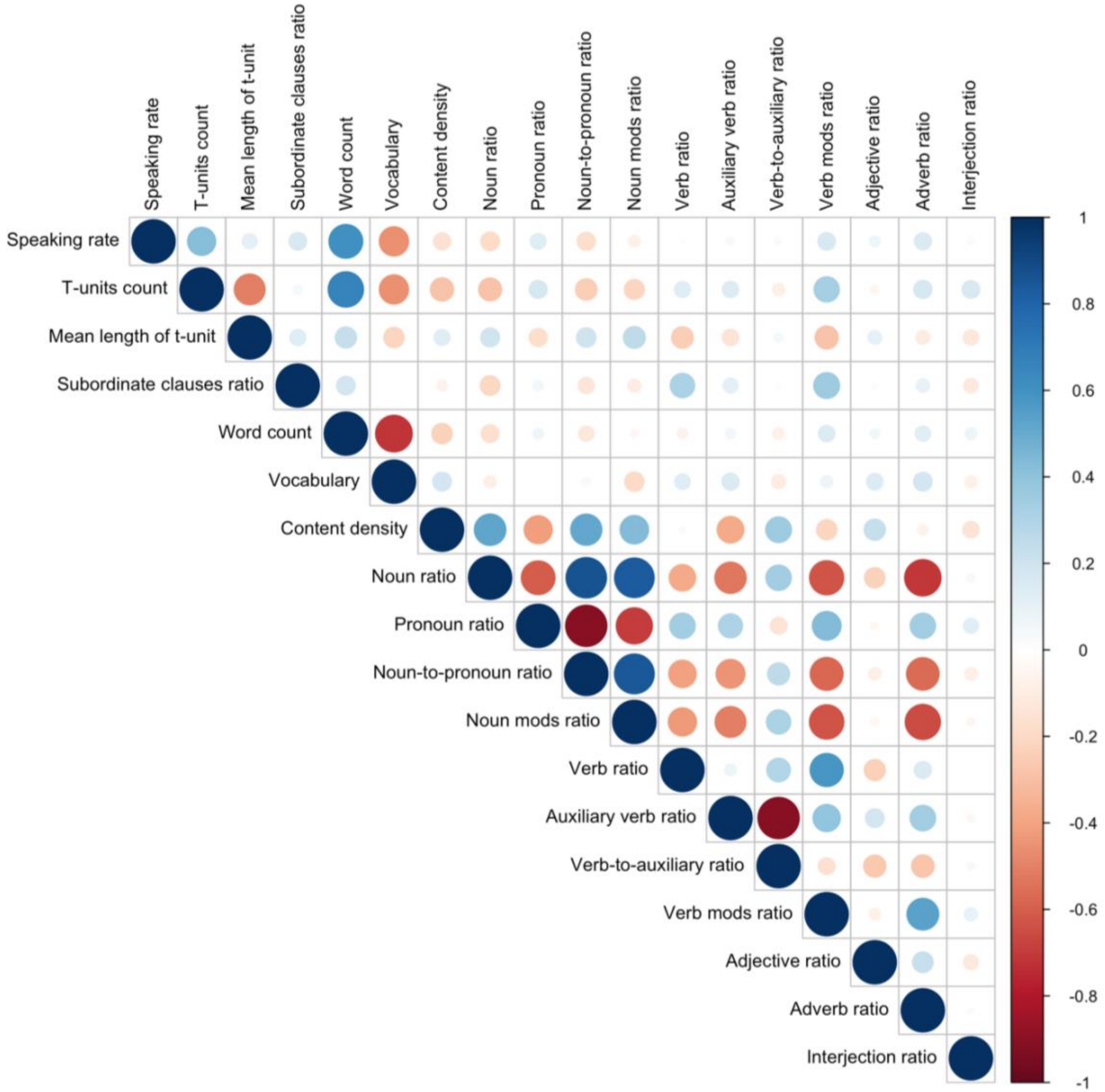
